# Boosting of the SARS-CoV-2-specific immune response after vaccination with single-dose Sputnik Light vaccine

**DOI:** 10.1101/2021.10.26.21265531

**Authors:** Alexey A. Komissarov, Inna V. Dolzhikova, Grigory A. Efimov, Denis Y. Logunov, Olga Mityaeva, Ivan A. Molodtsov, Nelli B. Naigovzina, Iuliia O. Peshkova, Dmitry V. Shcheblyakov, Pavel Volchkov, Elena Vasilieva

**Affiliations:** Clinical City Hospital named after I.V. Davydovsky, Moscow Department of Healthcare, 109240, 11/6 Yauzskaya str., Moscow, Russia; Federal State Budget Institution “National Research Centre for Epidemiology and Microbiology named after Honorary Academician N F Gamaleya” of the Ministry of Health of the Russian Federation, 123098, 18 Gamaleya str., Moscow, Russia; National Research Center for Hematology, 125167, 4a Novy Zykovsky proezd, Moscow, Russia; Genome Engineering lab, Moscow Institute of Physics and Technology, 141700, 9 Institutskiy per., Dolgoprudniy, Russia; A.I. Yevdokimov Moscow State University of Medicine and Dentistry, 127473, 20 Delegatskaya str., Moscow, Russia

**Author notes:** Corresponding author (A.A.K.).

## Abstract

Despite the measures taken worldwide, COVID-19 pandemic still progresses. While efficient antiviral drugs are not yet widely available, vaccination is the best option to control the infection rate. Although this option is obvious in case of COVID-19–naïve individuals, it is still unclear when individuals who have recovered from a previous SARS-CoV-2 infection should be vaccinated and whether the vaccination raises immune responses against the coronavirus and its novel variants. Here we measured the dynamics of the antibody and T-cell responses, as well as virus neutralizing activity (VNA) in serum against two SARS-CoV-2 variants, B.1.1.1 and B.1.617.2, among 84 individuals with different COVID-19 status who were vaccinated with Sputnik Light vaccine. We showed that vaccination of individuals previously exposed to the virus considerably boosts the existing immune response. In these individuals, RBD-specific IgG titers and VNA in serum were already elevated on the 7th day after vaccination, while COVID-19–naïve individuals developed the antibody response and VNA mainly 21 days post–vaccination. Additionally, we found a strong correlation between RBD-specific IgG titers and VNA in serum, and according to these data vaccination may be recommended if the RBD-specific IgG titers drop to 142.7 BAU/mL or below. In summary, the results of the study demonstrate that vaccination is beneficial both for COVID-19–naïve and recovered individuals, especially since it raises serum VNA against the B.1.617.2 variant – one of four the SARS-CoV-2 variants of concern.

## Introduction

As the new coronavirus disease 2019 (COVID-19) epidemic progresses, a growing number of individuals are becoming infected and cleared of its causative agent, the SARS-CoV-2 coronavirus, thus acquiring immune responses against this virus. Recent studies showed that acquired immunity protects to some extent from re-infection and severe disease [1-3]. However, the strength of the immune response varies considerably between individuals and is prone to decrease over time [4, 5]. Moreover, new SARS-CoV-2 variants have already been shown to escape from the immune responses developed as a result of infection with previous variants [6, 7]. While efficient antiviral drugs are not yet widely available, the best option to control the infection rate is vaccination against COVID-19. Although this option is obvious in case of COVID-19–naïve individuals, it is still unclear when individuals who have recovered from a previous SARS-CoV-2 infection should be vaccinated and to what extent the vaccination raises immune responses against the coronavirus and its novel variants in these individuals.

Recently, the single-component “Sputnik Light” vaccine, which represents the first component of the “Sputnik V” vaccine (recombinant human adenovirus serotype 26 bearing the gene of the SARS-CoV-2 spike protein [8]) was registered and approved for application in Russia (ClinicalTrials.gov Identifier: NCT04741061). This vaccine is considered as a promising boost when coronavirus-specific IgG titers in blood have decreased after two-dose vaccination or after recovery from COVID-19. In the current study, we focused on the effect of Sputnik Light on the latter population. We collected blood from the COVID-19–recovered individuals and from COVID-19–naïve ones prior to the Sputnik Light inoculation, followed by blood collection from the same individuals on days 7 and 21 post-vaccination, and compared in the collected specimens the dynamics of the antibody and T-cell responses. As well, we compared virus neutralizing activity in serum against two SARS-CoV-2 variants – B.1.1.1 and B.1.617.2.

## Materials and methods

### Ethics

This study was approved by the Moscow City Ethics Committee of the Research Institute of the Organization of Health and Healthcare Management and performed according to the Helsinki Declaration. All participants provided their written informed consent. This study is a part of a project that has been registered on ClinicalTrials.gov (Identifier: NCT04898140). After providing written informed consent, the individuals hand-filled a questionnaire containing information about their demographics, health, marital and social status, and self-estimated previous COVID-19 status or possible contacts with COVID-19 positive individuals.

### Blood collection and PBMC isolation

Peripheral blood was collected into two 8-mL VACUTAINER® CPT(tm) tubes with sodium citrate (BD, USA) and was processed within two hours after venipuncture. We isolated peripheral blood mononuclear cells (PBMC) according to the manufacturer’s standard protocol by centrifugation at 1,800–2,000*g* for 20 min with slow brake at room temperature (RT). After centrifugation, PBMC were collected into a 15-mL conical tube, washed twice with phosphate buffered saline (PBS, PanEco, Russia) with EDTA at 2 mM (PanEco, Russia), counted, and used for IFNγ ELISpot assay. PBMC with a viability level ≥70% were taken into the study. For serum isolation, peripheral blood was collected into S-Monovette 7.5-mL Z tubes (Sarstedt, Germany).

### SARS-CoV-2–specific antibodies and virus neutralizing activity in serum

Titers of the immunoglobulins G (IgGs) specific to the receptor-binding domain (RBD) of the SARS-CoV-2 spike (S) protein were analyzed in serum using the automated ARCHITECT i1000SR analyzer with compatible reagent kit (Abbott, USA) according to the manufacturer’s standard protocol. Values obtained were recalculated in BAU/mL in accordance with the WHO International Standard [9]; the IgG value equal to 7.2 BAU/mL was used as a seropositivity cutoff according to the manufacturer’s instructions. We evaluated the virus neutralizing activity (VNA) in serum from a microneutralization assay using B.1.1.1 (PMVL-1 (GISAID EPI_ISL_421275)) and B.1.617.2 (T19R G142D E156G F157del R158del L452R T478K D614G P681R D950N) SARS-CoV-2 variants in a 96-well plate and a 50% tissue culture infective dose (TCID50) of 100 as described in [10], with serum dilutions of 10, 20, 40, 80, 160, 320, 640, 1280, 2560, 5120, and 10,240 times.

### IFNγ ELISpot assay

We performed an IFNγ ELISpot assay using the Human IFNγ Single-Color ELISPOT kit (CTL; USA) with a 96-well nitrocellulose plate pre-coated with human IFNγ capture antibody according to the manufacturer’s protocol. Briefly, 3×10^5^ freshly isolated PBMC in serum-free CTL-test medium (CTL, USA), supplemented with Glutamax (ThermoFisher Scientific, USA) and penicillin/streptomycin (ThermoFisher Scientific, USA), were plated per well and incubated with SARS-CoV-2 PepTivator N or M or a mixture of S, S1, and S+ peptide pools (Miltenyi Biotec, Germany) at a final concentration of 1 μg/mL each at a final volume of 150 µL/well. Additionally, cells were incubated with media only (negative control) or phytohaemagglutinin (Paneco, Russia) at a final concentration of 10 µg/mL (positive control). Plates were incubated for 16–18 h at 37°C in 5% CO2 atmosphere. The plates were washed twice with PBS, then washed twice with PBS containing 0.05% Tween-20, and incubated with biotinylated anti-human IFNγ detection antibody for 2 h at RT. Plates were washed three times with PBS containing 0.05% Tween-20 followed by incubation with streptavidin-AP for 30 min at RT. We visualized spots by incubation with the substrate solution for 15 min at RT. The reaction was stopped by a gentle rinse with tap water. We air-dried plates overnight at RT and then counted spots using the automated spot counter CTL ImmunoSpot Analyzer and ImmunoSpot software (CTL; USA). Samples in which the negative control was greater than 10 spots and/or the positive control was less than 20 spots were considered as invalid. Positivity criteria for ELISpot were developed previously [11].

### Statistical analysis

Statistical analysis was performed with the Python3 programming language with *numpy, scipy* and *pandas* packages. The Mann–Whitney U test (two-sided) was used for comparing distributions of quantitative parameters between independent groups of individuals. The Wilcoxon signed-rank test (two-sided, including zero-differences in the ranking process and splitting the zero rank between positive and negative ones) was performed to assess the changes in the quantitative parameters between different time points for the same subject. To control for type I error, we calculated false discovery rate q-values using the Benjamin– Hochberg (BH) procedure and set a threshold of 0.05 to keep the positive false discovery rate below 5%. In all figures, for simplicity, we ranked p-values by significance levels using the following labels: 5.00e-02 < p-values are marked with ‘ns’; 1.00e-02 < p-values ≤ 5.00e-02 are marked with ‘*’; 1.00e-03 < p-values ≤ 1.00e-02 with ‘**’; 1.00e-04 < p-values ≤ 1.00e-03 with ‘***’; and p-values ≤ 1.00e-04 with ‘****’.

For the assessment of the different groups of subjects, a hierarchical cluster analysis using Ward variance minimization algorithm on z-normalized values for RBD-specific IgG levels at three time points for each subject was performed.

To select the optimal RBD-specific IgG titers for selection of VNA-positive patients, the binary classifier separating patients into groups with VNA >=20 and <20 for either B.1.1.1 or B.1.617.2 using RBD-specific IgG titers as a single input parameter was built and corresponding ROC curves were used for selection of optimal thresholds.

## Results

### Cohort characteristics

A total of 84 initially non-vaccinated Moscow residents were included in the study (**Table 1**). In the course of the study, participants were vaccinated with Sputnik Light vaccine and their blood was collected prior to the vaccination, as well as on days 7 and 21 after vaccine administration.

**Table 1.** General characteristics of the cohort

| Parameter | N = 84 |
| --- | --- |
| Age, years, median [IQR] | 45 [36, 54] |
| Females, N (%) | 65 (77.4%) |
| BMI, kg/m<sup>2</sup>, median [IQR] | 25.0 [22.1, 28.7] |
| Comorbidities, N (%): |  |
| Hypertension | 9 (10.7%) |
| Diabetes | 2 (2.4%) |
| Pulmonary diseases | 1 (1.2%) |
| Thyroid gland diseases | 3 (3.6%) |
| Gastritis | 6 (7.1%) |

BMI, body mass index.

### Changes in antibody and T-cell response levels

Serological testing of the participants on the day of their inclusion in the study revealed that 44 (52.4%) individuals were seropositive for the virus-specific immunoglobulins G (IgGs) (**Figure 1A**). Among them, 36 (81.8%) individuals also demonstrated the presence of SARS-CoV-2–specific T cells in peripheral blood (**Figure 1B**). These data, taken together with the self-reported COVID-19 cases in this group, indicated that these individuals had been previously exposed to SARS-CoV-2.

**Figure 1.**
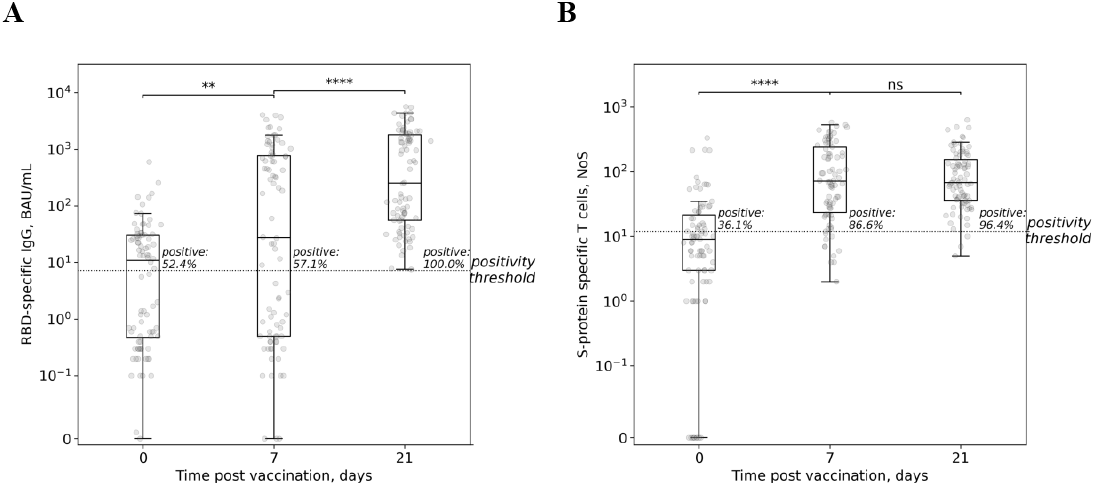
Dynamics of the antibody and T-cell responses. Titers of the IgGs specific to the receptor binding domain (RBD) of the SARS-CoV-2 spike (S) protein (A) and frequencies of the S-protein specific T cells in peripheral blood (B) were estimated at the indicated days post-vaccination and represented as standard box-and-whiskers diagrams with individual values represented by dots. NoS, number of spots estimated by ELISpot (see the Materials and Methods section for details).

Within the observational period among the cohort, there was a constant increase in the titers of IgGs specific to the receptor binding domain (RBD) of the coronavirus spike (S) protein (**Figure 1A**). Accordingly, the fraction of seropositive individuals also increased from 52.4% to 57.1% on the 7th day, and to 100% on the 21st day post-vaccination. The results of the IFNγ ELISpot demonstrated that the T-cell response developed faster than the antibody one. Frequencies of peripheral blood T cells specific to coronavirus S-protein had already increased on the 7th day post-vaccination and didn’t change significantly on the 21st day (**Figure 1B**), with the fraction of individuals positive for T-cell response being 36.1%, 86.6%, and 96.4% on days 0, 7, and 21 post vaccination, respectively. Meanwhile, as expected, the fractions of individuals who demonstrated the presence in peripheral blood of T cells specific to membrane (M) and nucleocapsid (N) SARS-CoV-2 proteins didn’t change until the end of the observation period (**Supplementary Figure S1**).

### Participant clusterization by the types of immune response dynamics

To find the main patterns of the response to the vaccination, all participants were clusterized according to the observed dynamics in RBD-specific IgG titers. For this purpose, we used the Ward variance minimization algorithm in order to discriminate these patterns in an unbiased way. Three clusters were identified (**Figure 2 and Supplementary Figure S2**). Clusters 1 (*n* = 42) and 2 (*n* = 4) were composed of the individuals who were seropositive at the time of their inclusion in the study, except for two individuals from cluster 1 with IgG levels equal to 5.6 and 6.2 BAU/mL, which fall below the seropositivity cutoff of 7.2 BAU/mL according to the serological test manufacturer. Meanwhile, cluster 3 (*n* = 38) was composed of the seronegative individuals only, with the highest IgG level equal to 2.0 BAU/mL. No statistically significant differences between clusters in available clinical data were found.

**Figure 2.**
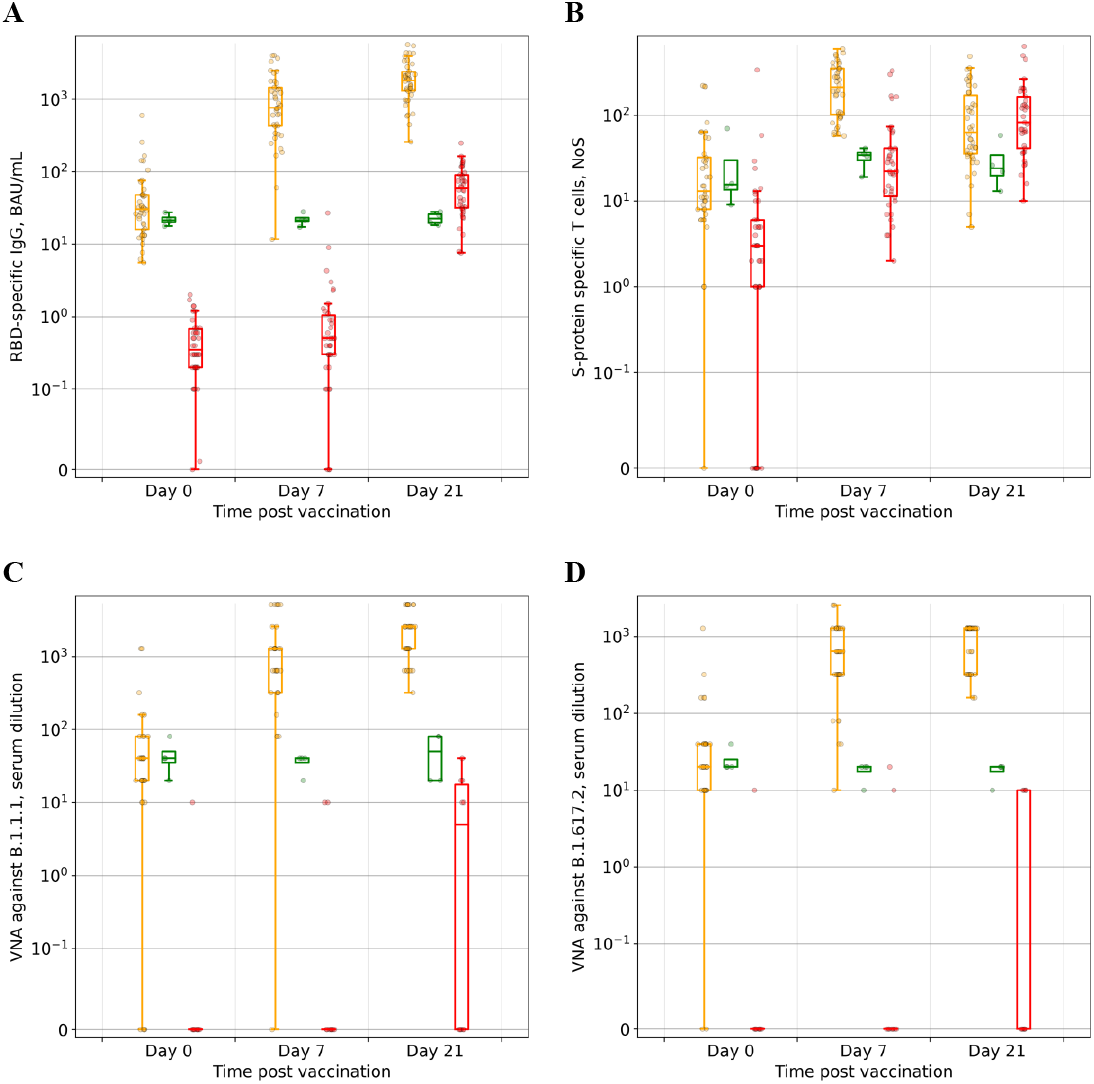
Characteristics of the different clusters of participants. For each cluster, receptor-binding domain (RBD) specific IgG titers (A), frequencies of S-protein–specific T cells (B), as well as virus neutralizing activity (VNA) in serum against B.1.1.1 (C) and B.1.617.2 (D) SARS-CoV-2 variants, were estimated as described in the Materials and Methods section and represented as standard box-and-whiskers diagrams with individual values represented by dots. NoS, number of spots estimated by ELISpot. Orange corresponds to Cluster 1, green to Cluster 2, red to Cluster 3.

All individuals from cluster 3 demonstrated no increase in IgG levels on the 7th, but the titers were significantly elevated on the 21st day post-vaccination (**Figure 2A and Supplementary Figure S3A**). At the same time, we found a monotonous increase in the frequencies of S-protein specific T cells throughout the observation period (**Figure 2B and Supplementary Figure S3B**).

Different results were observed in individuals previously exposed to SARS-CoV-2. Prior to the vaccination, clusters 1 and 2 were characterized by the same values of IgG titers and frequencies of S-protein–specific T cells. However, in the case of cluster 1, IgG titers had already increased considerably on the 7th day post-vaccination but were only slightly elevated on the 21st day. Frequencies of S-protein–specific T cells in peripheral blood also increased considerably 7 days post vaccination, but then dropped on the 21st day, reaching the same value as for cluster 3 (composed of the SARS-CoV-2–naïve individuals).

In contrast to cluster 1, individuals comprising cluster 2 demonstrated no changes in either IgG titers or frequencies of S-protein–specific T cells throughout the observation period. On the 21st day post-vaccination, both parameters were significantly lower than those for individuals without previous SARS-CoV-2 exposure (cluster 3) (**Figure 2A, B and Supplementary Figure S3A, B**).

### Virus neutralizing activity in serum against different SARS-CoV-2 variants

For a group of individuals from the cohort (47 participants), we analyzed the virus neutralizing activity (VNA) in serum against two SARS-CoV-2 variants - B.1.1.1 and

B.1.617.2. For this purpose, we used the microneutralization assay with different serum dilutions. Almost all individuals with previous SARS-CoV-2 exposure (those in clusters 1 and 2) demonstrated the presence of VNA against both variants even prior to the vaccination, with the values being indistinguishable (**Figure 2C, D and Supplementary Figure S3C, D**). Further, in case of cluster 1, VNA increased and reached the maximum already on the 7th day post-vaccination, while VNA in serum of individuals in cluster 2 didn’t change for either virus variant until the end of the observation period. For SARS-CoV-2–naïve individuals from cluster 3, VNA in serum against both variants appeared mainly on the 21st day.

It should be noted that VNA values against both SARS-CoV-2 variants at each time point were proportional (**Figure 3A**); however, VNA against the B.1.617.2 variant was approximately two times lower than that against the B.1.1.1 variant (**Figure 3B**).

**Figure 3.**
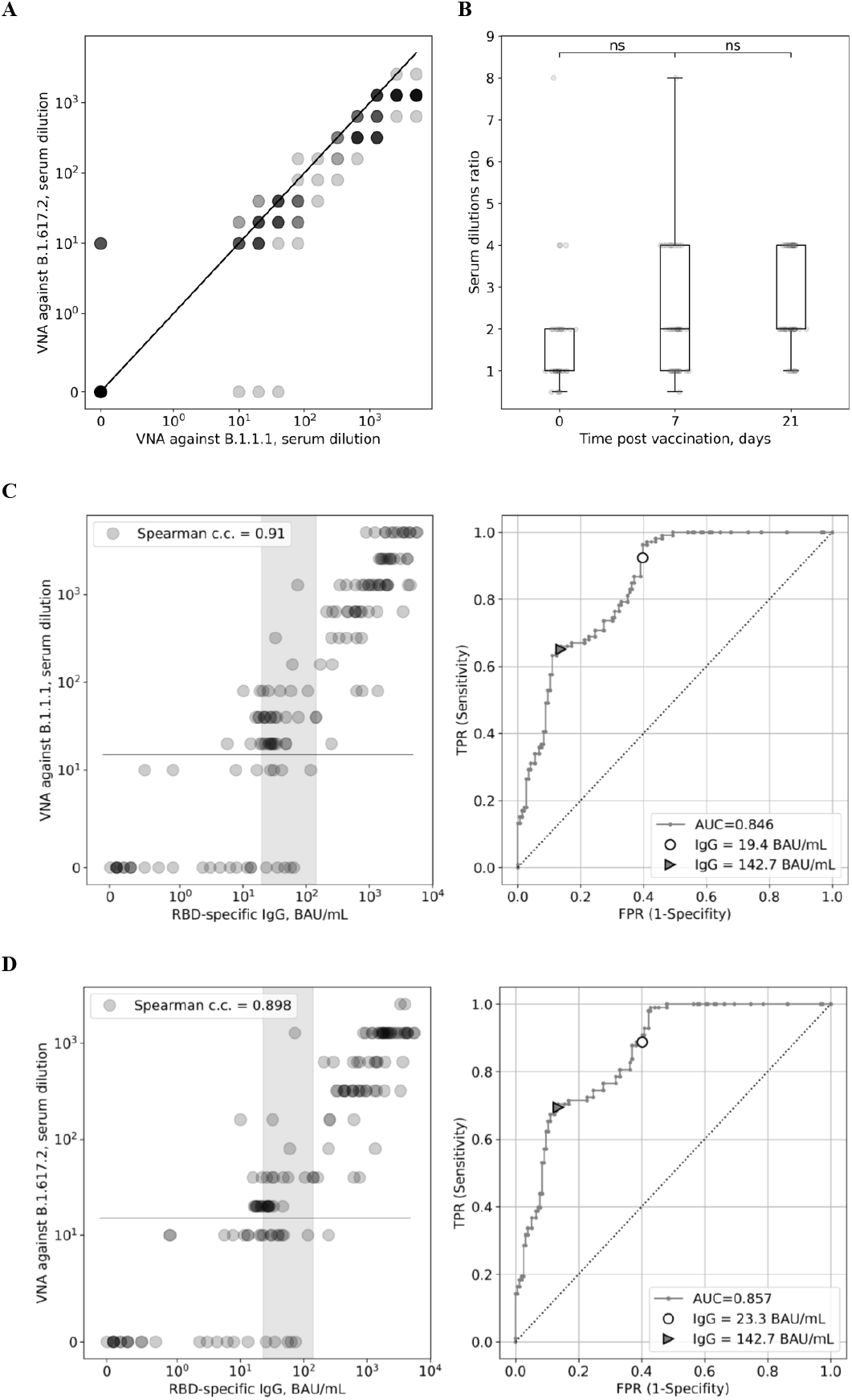
Analysis of the virus neutralizing activity in serum against B.1.1.1 and B.1.617.2 SARS-CoV-2 variants. **A:** Dot plots for comparison between virus neutralizing activities (VNA) is serum against B.1.1.1 and against B.1.617.2 for all patients at all time points. **B:** Boxplots for ratio of VNA against B.1.1.1 to VNA against B.1.617.2 for all patients at different time points with both VNA against B.1.1.1 and VNA against B.1.617.2 above 0. **C: Left:** dot plot for pairwise comparisons of Abbott IgG and VNA against B.1.1.1 for all patients at all time points; zone between markers from the right panel is shown in grey. **Right:** ROC curve for binary classifier separating patients into groups with VNA against B.1.1.1 >=20 and <20 using RBD specific IgG titers as a single input parameter. Two potential optimum thresholds are shown with a marker. **D: Left:** dot plot for pairwise comparisons of Abbott IgG and VNA against B.1.617.2 for all patients at all time points; zone between markers from the right panel is shown in grey. **Right:** ROC curve for binary classifier separating patients into groups with VNA against B.1.617.2 >=20 and <20 using RBD specific IgG titers as a single input parameter. Two potential optimum thresholds are shown with a marker.

For each of the tested SARS-CoV-2 variants, we found a strong correlation between RBD-specific IgG titers and VNA in serum (**Figure 3C, D**). Accordingly, these IgG titers can be used as a predictor of the presence of VNA in serum. To minimize the impact of possible false positive results on prediction, we set up the second serum dilution used in the study (20 times, see the Materials and Methods section for details) as a threshold for the presence of VNA. Analysis of the corresponding ROC curves revealed that the highest sensitivity of the prediction was achieved at 19.4 and 23.3 BAU/mL for the B.1.1.1 and B.1.617.2 variants, respectively. However, the optimal specificity was achieved at 142.7 BAU/mL for both SARS-CoV-2 variants. Meanwhile, individuals having RBD -specific IgG levels in a range 19.4(23.3)–142.7 BAU/mL were characterized by relatively low levels of VNA in serum or even by the absence of the neutralization activity.

## Discussion

In the present study, we analyzed the effect of the Sputnik Light vaccine administration on the anti-SARS-CoV-2 immune response in individuals with different COVID-19 statuses. For this purpose, we collected blood from participants prior to vaccination, and at time points after vaccine administration. We analyzed the development of (i) immunoglobulins G (IgGs) specific to the receptor-binding domain (RBD) of the coronavirus spike (S) protein, (ii) S-protein–specific T cells in peripheral blood, and (iii) virus neutralizing activity (VNA) in serum against two SARS-CoV-2 variants - B.1.1.1 and B.1.617.2.

Upon vaccination with Sputnik Light, we observed different dynamics of both antibody and T-cell responses depending on the previous SARS-CoV-2 infection status of the tested individuals. In accordance with published data [8], in COVID-19–naïve individuals, coronavirus-specific IgGs appeared largely on the 21st day post-vaccination, and similar results were obtained for VNA against both coronavirus variants. In contrast, S-protein– specific T cells appeared in peripheral blood already on the 7th day, and their number further increased on the 21st day post-vaccination. Meanwhile, already prior to vaccine administration, 8 of 40 seronegative individuals were characterized by the presence of S-protein–specific T cells, with 6 among them being also positive for T cells specific to membrane and nucleocapsid proteins of SARS-CoV-2. The presence of SARS-CoV-2– specific T cells in these seronegative individuals might be explained by previously asymptomatic COVID-19, which has been shown to be associated with lack of antibody response or a rapidly decreasing one [12, 13], or these T cells might have developed as a result of a previous infection with the “common cold” coronaviruses and are cross-reactive to SARS-CoV-2 [14, 15].

As expected, individuals infected with SARS-CoV-2 prior to vaccination were characterized by the presence of the SARS-CoV-2–specific IgGs and T cells, as well as of the VNA in serum, before the vaccination. For the vast majority of these individuals, all these parameters had already increased considerably on the 7th day post-vaccination, thus indicating that for the recovered persons Sputnik Light served as an effective booster. Similar results were obtained for the Pfizer (BNT162b2) and Moderna (mRNA-1273) mRNA vaccines [16], as well as for another single-dose adenovirus-vectored vaccine, ChAdOx1 nCoV-19 [17]. However, among seropositive individuals only four persons (11% of the seropositive group) did not respond to the vaccination with Sputnik Light, as evidenced by the lack of increase in anti-SARS-CoV-2 IgG titers, peripheral blood T cells, and VNA in serum. While the reasons for this lack have yet to be understood, the fraction of non-responders is rather small and doesn’t compromise the general efficacy of vaccination among COVID-19–recovered individuals.

Recent studies have shown that IgGs specific to the coronavirus S-protein, particularly to its RBD portion, also demonstrate neutralizing activity against the virus [18-20]. Similar results were obtained in our study: we found a strong correlation between RBD-specific IgG titers and VNA in serum. This correlation was especially pronounced in the case of COVID-19– naïve individuals: VNA in their serum appeared simultaneously with the appearance of the RBD-specific IgGs evaluated on day 21 post-vaccination. It is noteworthy that by the end of the observation period all initially sero-negative individuals had become seropositive; however, not all of them developed VNA in serum. It is likely that this discrepancy originates from the individual features of the immune response kinetics, and the discrepancy will be leveled at distant time points post-vaccination. For COVID-19–recovered individuals, however, VNA was detected in all serum samples and it had already increased on day 7 post vaccination.

On the basis of evidence from clinical trials and convalescent cohort studies, it was recently found that it is neutralizing antibodies that mainly correlated with protection from SARS-CoV-2 infection and from the severe disease form [21]. In our study, we found that the presence of VNA in serum can be reliably estimated on the basis of the RBD-specific IgG titers. For both the B.1.1.1 and B.1.617.2 SARS-CoV-2 variants, individuals with IgG levels higher than 142.7 BAU/mL demonstrated the highest VNA and therefore are likely characterized by the highest protectivity. These data are in agreement with another study in which the same serological test was used [20].

In accordance with published data [22], we found that VNA developed against the B.1.617.2 SARS-CoV-2 variant after vaccination with Sputnik Light was approximately two times lower than that against the B.1.1.1 variant. Nevertheless, we showed that, depending on the COVID-19 status of the individual, vaccination promotes the formation of or significantly increases the VNA against the B.1.617.2 variant, one of four the SARS-CoV-2 variants of concern, detected in Russia and many other countries (https://www.who.int/en/activities/tracking-SARS-CoV-2-variants).

Taken together, our results showed that vaccination with Sputnik Light in cases of individuals previously exposed to the virus considerably boosts the existing immune response against the virus. In these individuals, RBD-specific IgG titers, S-protein–specific T cells, and VNA in serum were already elevated on the 7th day after vaccination, in contrast to the COVID-19– naïve individuals, who developed the antibody response and VNA in serum mainly 21 days post–vaccination. We found a strong correlation between RBD-specific IgG titers and VNA in serum, and according to these data vaccination may be recommended if the RBD-specific IgG titers drop to 142.7 BAU/mL or below. In summary, the results of the study demonstrate that vaccination is beneficial both for COVID-19–naïve and recovered individuals, especially since it raises serum VNA against the B.1.617.2 variant, and Sputnik Light can be efficiently used for this purpose.

## Data Availability

All data produced in the present work are contained in the manuscript and supplementary files

## Acknowledgements

The authors would like to thank Dr. Leonid Margolis (Eunice Kennedy Shriver National Institute of Child Health and Human Development, National Institutes of Health, Bethesda, MD, USA) for his valuable comments and suggestions during experimental design, discussion of the results and manuscript preparation, and Dr. Barry Alpher for assistance in editing and improving the English style of the manuscript. The authors also thank the Moscow Department of Healthcare for the help in organization of the study.

## Supplementary materials

**Figure S1.**
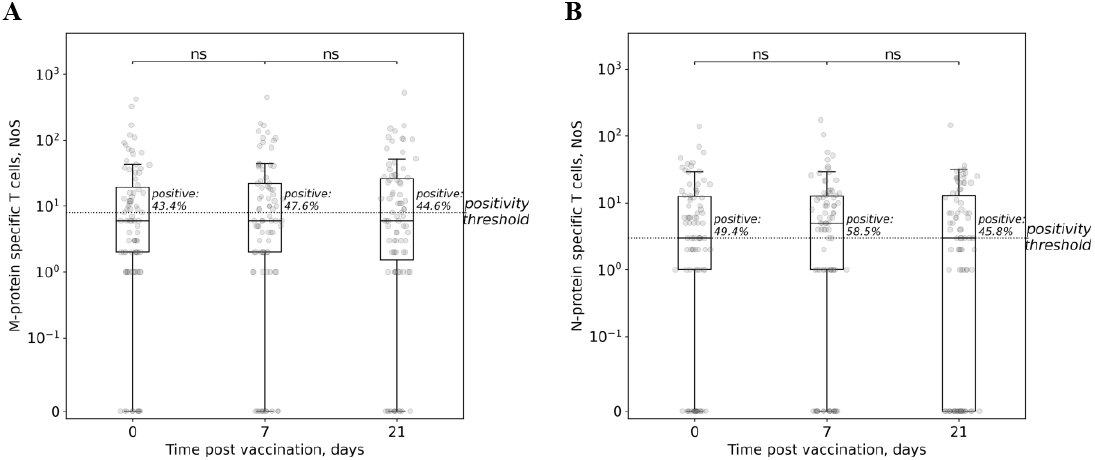
Dynamics of the T-cell response in the cohort. Frequencies of the M-(A) and N-protein (B) specific T-cells in peripheral blood were estimated at the indicated days post vaccination and represented as standard box-and-whiskers diagrams with values represented by dots.

**Figure S2.**
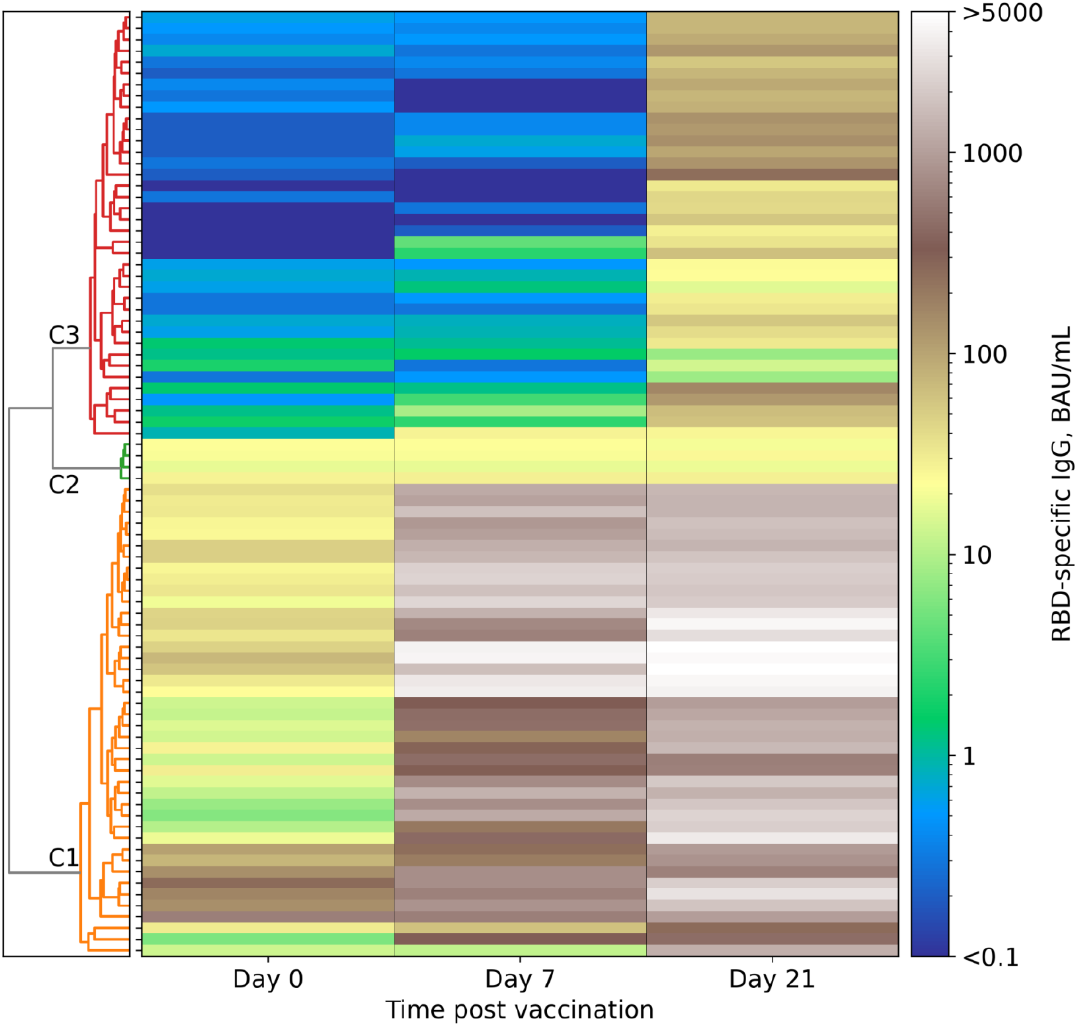
Clusterization of the participants by the dynamics of the antibody response. RBD-specific IgG titers were estimated as described in the Materials and Methods section and were represented as a heatmap. Next, participants were clusterized by the changes in the IgG titers with the use of the Ward variance minimization algorithm. Clusters found are indicated as C1, C2, and C3.

**Figure S3.**
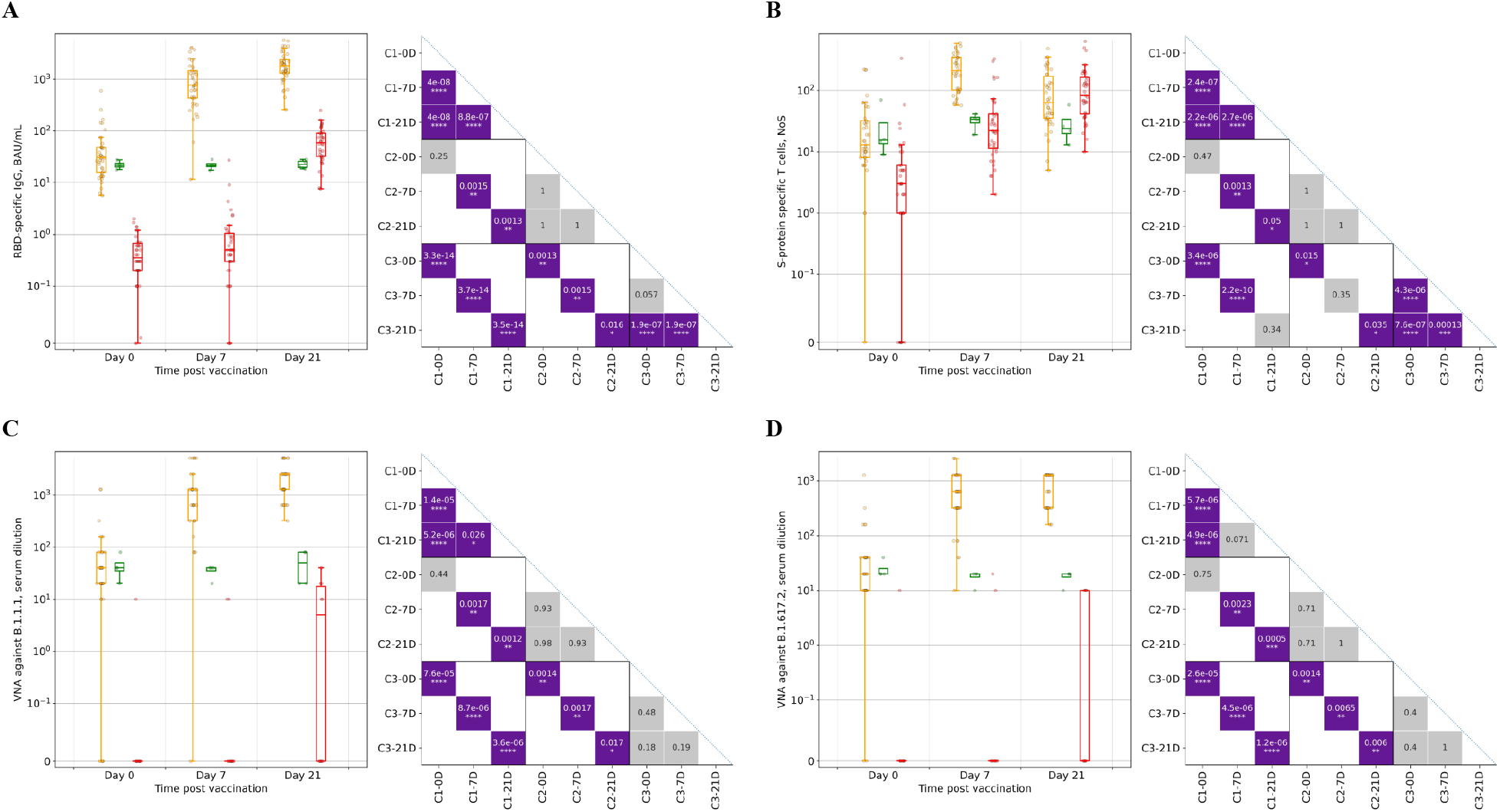
Characteristics of the different clusters of participants. For each cluster, RBD-specific IgG titers (A), frequencies of S-protein–specific T cells (B), as well as virus neutralizing activity (VNA) in serum against B.1.1.1 (C) or B.1.617.2 (D) SARS-CoV-2 are shown. Variants were estimated as described in the Materials and Methods section. For each panel, at left are boxplots for different clusters at indicated time points; orange corresponds to Cluster 1, green to Cluster 2, red to Cluster 3. At right are shown *p*-values for comparison between different clusters (Cluster, 1, 2, and 3 are designated as C1, C2, and C3, respectively) and/or different time points (0, 7, and 21 days post vaccination are designated as 0D, 7D, and 21D, respectively). For comparisons between different time points within same clusters, *p*-values from the Wilcoxon paired test are given; for comparisons between different clusters at the same time point, *p*-values from the Mann-Whitney test are given. We corrected all *p*-values corrected within each family of comparisons using the Benjamini-Hochberg method. Statistically significant differences (*p*-value < 0.05) are highlighted with purple.

